# Disparities in outcomes among patients diagnosed with cancer associated with emergency department visits

**DOI:** 10.1101/2021.03.03.21252826

**Authors:** Nicholas Pettit, Elisa Sarmiento, Jeffrey Kline

**Affiliations:** Indiana University, School of Medicine. Department of Emergency Medicine

## Abstract

**Importance:** Diagnosis of cancer in the emergency department (ED) may be associated with poor outcomes, related to socioeconomic (SES) disparities, however data are limited.

**Objective:** To examine the morality and associated disparities for cancer diagnoses made less than six months after an ED visit.

**Design:** This study is case-control analysis of the Indiana State Department of Health Cancer Registry, and the Indiana Network for Patient Care. First time diagnoses of ICD-“cancer” appearing in the registry between January 2013 and December 2017 were included. Cases were patients who had an ED visit in the 6 months before their cancer diagnosis; controls had no recent ED visits.

**Main Outcome(s) and Measure(s):** Primary outcome was mortality, comparing ED-associated mortality to non-ED-associated. Secondary outcomes include SES and demographic disparities.

**Results:** 134,761 first-time cancer patients were identified, including 15,432 (11.5%) cases. In cases and controls, the mean age was same at 65 and the Charlson Comorbidity Index (CCI) was the same at 2.0 in both groups. More of the ED cohort were Black than the non-ED cohort (12.4% vs 7.4%, P<.0001, Chi Square) and more were low income (36.4%. vs 29.3%). The top 3 ED-associated cancer diagnoses were lung (18.4%), breast (8.9%), and colorectal cancer (8.9%), vs. the non-ED cohort were breast (17%), lung (14.9%), and prostate cancer (10.1%). Regardless of type, all ED-associated cancers had an over three-fold higher mortality, with cumulative death rate of 32.9% for cases vs 9.0% for controls (P<.0001) over the entire study period. Regression analysis predicting mortality, clustering by city, controlling for age, gender, race, SES, drug/alcohol/tobacco use, and CCI score, produced an odds ratio of 4.12 (95% CI 3.72-4.56 for ED associated cancers).

**Conclusion and Relevance:** This study found that an ED visit within 6 months prior to the first time of ICD-coded cancer is associated with Black race, low income and an overall three-fold increased risk of death. The mortality rates for ED-associated cancers are uniformly worse for all cancer types. These data suggest that additional work is needed in order to reduce disparities among ED-associated cancer diagnoses, including increased surveillance and improved transitions of care.

**Key Points:** *Question:* Do patients diagnosed with cancer, shortly after an emergency department (ED) visit have worsened outcomes than patients with diagnoses not associated with an ED visit.

*Findings:* In this retrospective, epidemiological assessment, 134,761 patients were diagnosed with cancer, 11.5% (15,432) were seen in the ED within 6 months prior to their diagnosis. They had increased mortality, associated with racial and socioeconomic disparities.

*Meaning:* These findings imply an urgent need for improved transitions of care for minority and low-income patients with suspected cancer in the ED.

## Introduction

Cancer is a leading cause of death worldwide.^1^ The number of cancer survivors continues to grow in the face of improving therapies and earlier detection, as well as declining age-standardized incidence rates in men and stable in women.^2^ Improved detection modalities is one reason for these improvements. Longitudinal outcome studies have suggested that each week added to the time to initial cancer treatment is associated with a 1-3% absolute increased risk of mortality with breast, lung, renal, and pancreatic cancers.^3^ National guidelines exist for cancer screening that are relevant to emergency care. For example, the U.S. Preventive Task Force recommends yearly lung cancer screening with low-dose CT for chronic, heavy smokers, between the ages of 55-80 years old. However, only a minority of patients are appropriately screened as outpatients (4% in the setting of lung cancer).^4^ This low frequency contrasts with the high public enthusiasm for screening (87%).^5^

Although poorly quantified, preliminary data suggest that many patients are diagnosed with cancer during an ED visit.^6^ Because patients who have cancer diagnosed during ED presentations may have multiple inequities in care (lack of insurance, no primary care, and possibly higher smoking rates), their outcomes may be worse. However, since the literature is sparse, and mostly retrospective, little is absolutely known about the outcomes of patients with emergently diagnosed cancer.

In this work, we use data linkage and case-control methodology to characterize the demographics, phenotypes, and outcomes of patients that have emergency department-associated cancer diagnoses. We used a statewide database to examine the rate of recent visit--defined arbitrarily as within six months--to an ED in patients with newly diagnosed cancer with the inference that the recent visit was the touchpoint where the initial clinical information suggested a possible new cancer diagnosis. We hypothesized that ED-associated cancer diagnoses will suffer higher mortality as compared to non-ED-associated cancer diagnoses.

## Design and Methods

### Summary of Data Extraction

This was a retrospective study using data from the Indiana State Department of Health (ISDH) Cancer Registry, the Indiana Network for Patient Care (INPC), supplemented with data from the electronic health records (EHR) of two major hospitals in the greater Indianapolis area.

Patients with their first cancer diagnosis between January 2013 and December 2017 were identified using the ISDH Cancer Registry. To determine presence or absence of a recent ED visit, clinical features and outcomes, patients in the ISDH Cancer registry were co-identified in the INPC research database, which represents one of the largest health information exchanges in the country. Cases were defined as patients with ED visits in the 6 months before their cancer diagnosis were and controls were all others. The index date for all patients was set as the date of initial cancer diagnosis according to the first posted cancer-defining ICD code. The ICD Oncology Topography codes, the cancer primary site, as well as patient zip code and insurance used at the time of diagnosis were extracted from the ISDH Cancer Registry.

Using unique identifiers for data linkage, demographics and clinical data, comorbid ICD codes were downloaded from the INPC and supplemented with cancer registry data. The Charlson Comorbidity Index (CCI) at time of index was calculated using diagnoses within the year prior to index. Social history (tobacco use, drug use, and alcohol use) was also identified using ICD codes. Tobacco use was supplemented with data available from local hospital electronic health records. Vital status and patient mortality was extracted using Social Security Administration (SSA) death data linked to the INPC. Where SSA data was missing, death data was extracted from the INPC and the ISDH Cancer Registry.

### Statistics

Patient demographics and characteristics were compared between those patients who did not have an ED visit within six months prior to cancer diagnosis (controls) and those patients who did (cases). To test for differences between groups, the Chi-square test or 95% confidence interval for differences in proportions were used for bivariate variables, and the Wilcoxon test for continuous variables. Adjusted odds ratios from logistic regression containing the following dependent variables (age, gender, race, SES, drug use, alcohol use, tobacco use, and the Charlson comorbidity index)was used to determine the association of ED diagnosis (cases) with minority status, low SES status and death compared with non-ED diagnosis (controls). A generalized estimating equation using the GENMOD procedure was used to adjust for clustering at the city level via zip code. We performed statistical analyses using SAS version 9.4 (SAS Institute, Cary, NC).

## Results

### Comparing the two populations

Table 1 gives the demographics and characteristics of the patients involved in this study, separating patients into controls, those with no-associated ED visits in prior 6 months and compares those to case patients with an associated ED visit within 6 months from the time of cancer diagnosis. In total, 134,761 patients were identified to be diagnosed with cancer within the Indiana Cancer Registry, with 15,432 (11.5%) of the patients having a recorded ED visit within 6 months of their diagnosis of cancer. First time cancer diagnoses were identified during the 2013-2017 study period but mortality was included up to 2019.

**Table 1.**
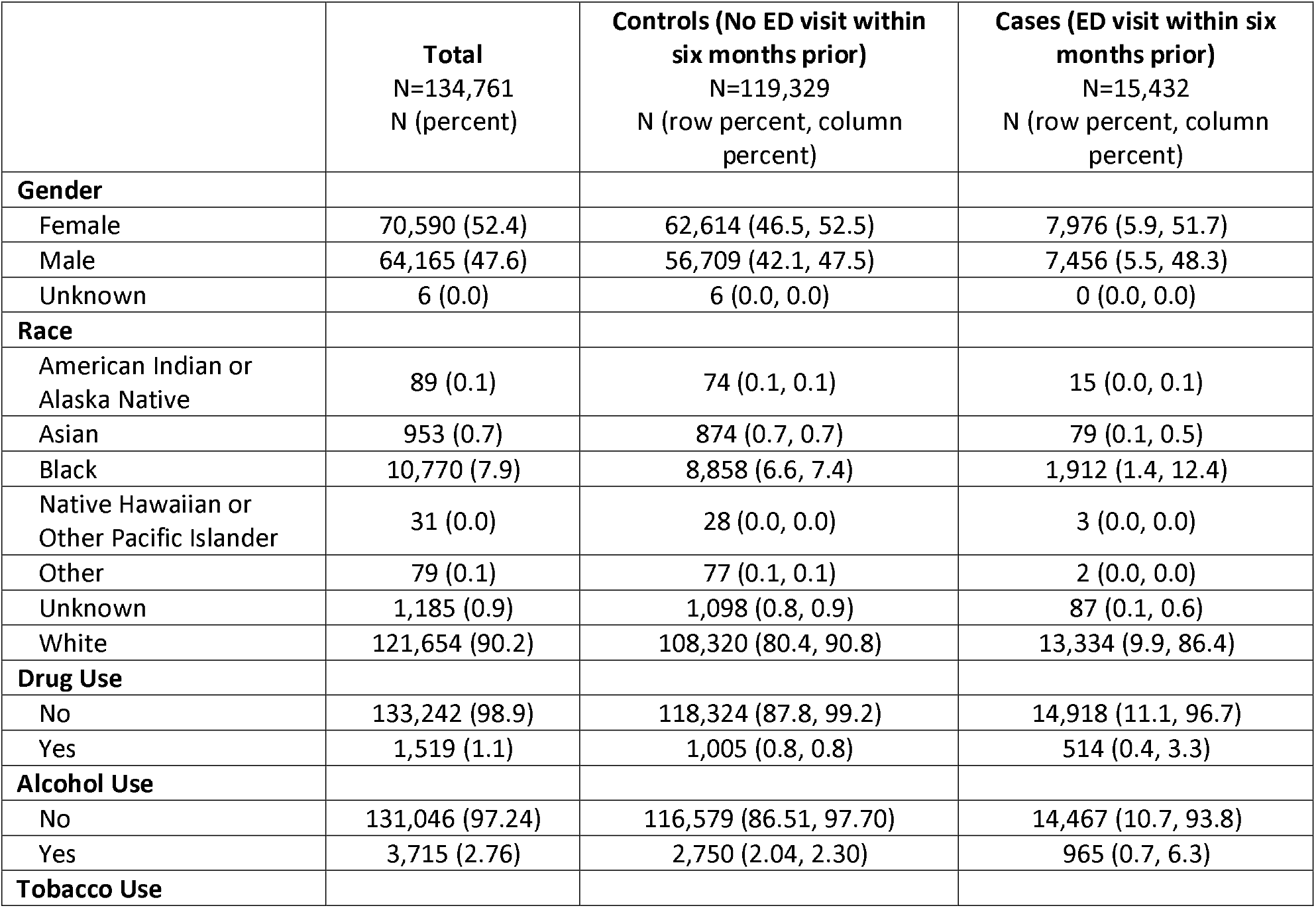
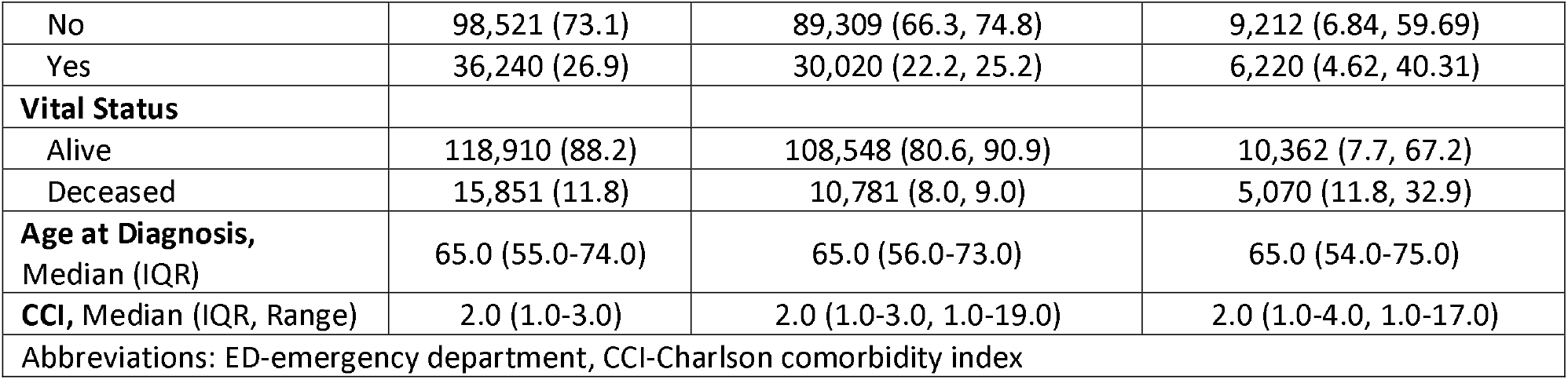
Characteristics of the study population

Pertinent differences included the finding that African Americans comprised a significantly greater proportion of cases (12.4% versus 7.4, 95% CI for difference of 5.0%= 2.3 to 6.4%). Furthermore, cases had higher rates of drug, alcohol, and tobacco use than controls at 3.3% vs 0.8% (95% CI for 2.5% = 0.20-0.25) for drug use, 6.3% vs 2.3% for alcohol use (95% CI For 4.0% = 0.14-0.16), and 40.3% vs 25.2% for tobacco use (95% CI for 15.1% = 0.07-0.08) In terms of primary outcome, cases had a significantly higher mortality rate of 32.8% versus 9.0% (95% CI for 23.8% = 0.23-0.24) within a median 116 days for cases and 268 days for controls.

### Top cancers per cohort

Table 2 lists the top 10 malignancies for both cohorts in order of decreasing frequency. The values within each column for frequencies gives two percentages, with the first percentage the overall percentage and the second being the percentage just within controls or cases. For controls, the top 3 cancers in decreasing frequency are breast (18.1%), lung (14.4%), and prostate (10.1%). Comparatively the most frequently ED-associated cancers were lung (18.4%), breast (8.9%), and colon/rectal (8.9%).

**Table 2.**
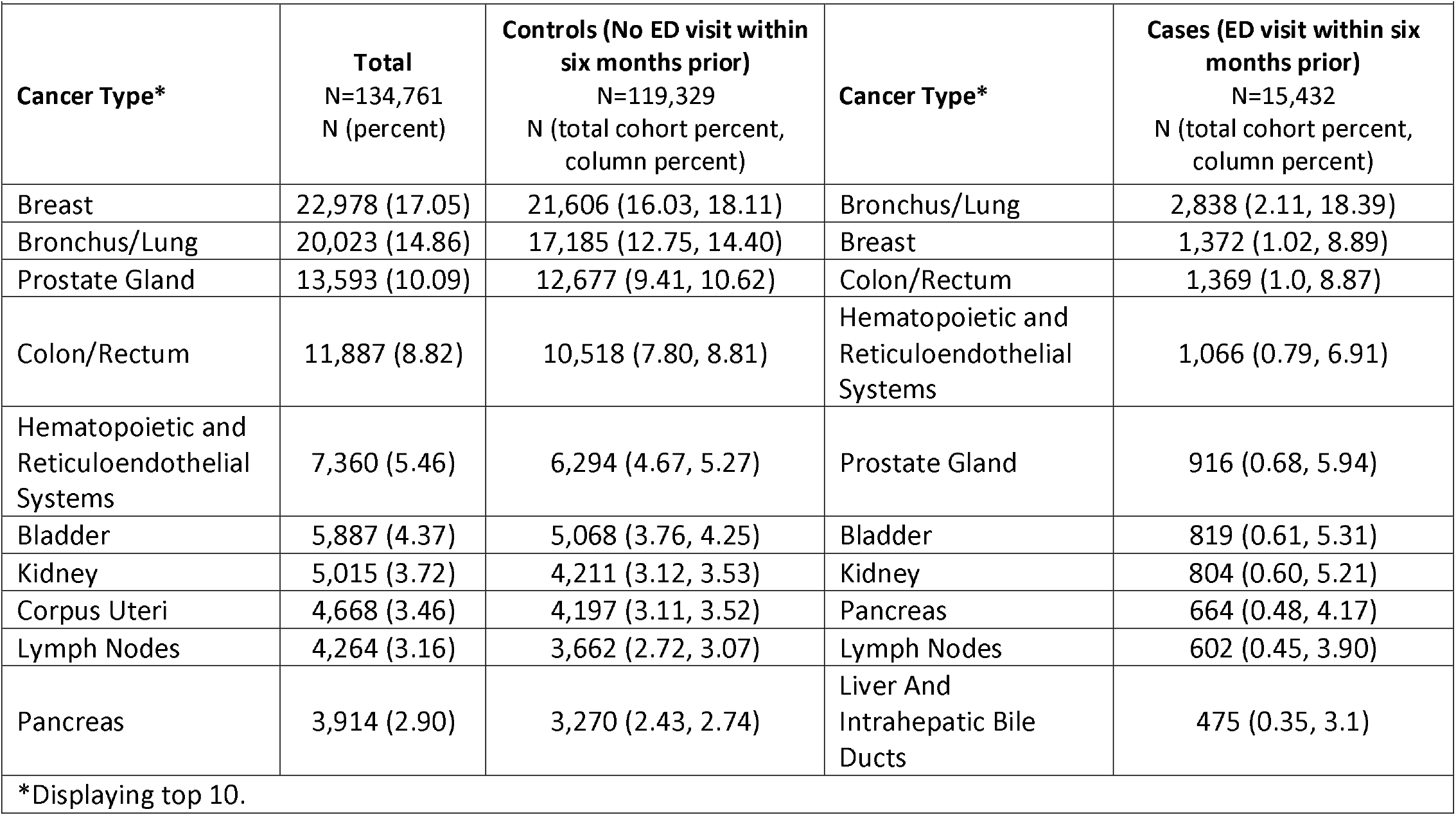
Frequency of cancers among controls and cases.

| **Cancer Type\*** | **Total**<br>N=134,761<br>N (percent) | **Controls (No ED visit within six months prior)**<br>N=119,329<br>N (total cohort percent, column percent) | **Cancer Type\*** | **Cases (ED visit within six months prior)**<br>N=15,432<br>N (total cohort percent, column percent) |
| --- | --- | --- | --- | --- |
| Breast | 22,978 (17.05) | 21,606 (16.03, 18.11) | Bronchus/Lung | 2,838 (2.11, 18.39) |
| Bronchus/Lung | 20,023 (14.86) | 17,185 (12.75, 14.40) | Breast | 1,372 (1.02, 8.89) |
| Prostate Gland | 13,593 (10.09) | 12,677 (9.41, 10.62) | Colon/Rectum | 1,369 (1.0, 8.87) |
| Colon/Rectum | 11,887 (8.82) | 10,518 (7.80, 8.81) | Hematopoietic and Reticuloendothelial Systems | 1,066 (0.79, 6.91) |
| Hematopoietic and Reticuloendothelial Systems | 7,360 (5.46) | 6,294 (4.67, 5.27) | Prostate Gland | 916 (0.68, 5.94) |
| Bladder | 5,887 (4.37) | 5,068 (3.76, 4.25) | Bladder | 819 (0.61, 5.31) |
| Kidney | 5,015 (3.72) | 4,211 (3.12, 3.53) | Kidney | 804 (0.60, 5.21) |
| Corpus Uteri | 4,668 (3.46) | 4,197 (3.11, 3.52) | Pancreas | 664 (0.48, 4.17) |
| Lymph Nodes | 4,264 (3.16) | 3,662 (2.72, 3.07) | Lymph Nodes | 602 (0.45, 3.90) |
| Pancreas | 3,914 (2.90) | 3,270 (2.43, 2.74) | Liver And Intrahepatic Bile Ducts | 475 (0.35, 3.1) |

\*Displaying top 10.

### Comparing Mortality Between Cancers

Figure 1 (and supplemental Table 1) compares the cumulative mortality rates for 9 cancer diagnoses during the 5-year study period. The percentage of mortality for all nine cancer types were consistently higher in cases compared with controls. A concerning 45% relative difference exists between cases and controls for pancreatic cancer. Figure 2 shows Kaplan Meier survival curves for these nine cancers and demonstrates a consistently worse survival for cases versus controls up until 5 years of follow-up (P<0.001 log-rank statistic).

**Figure 1.**
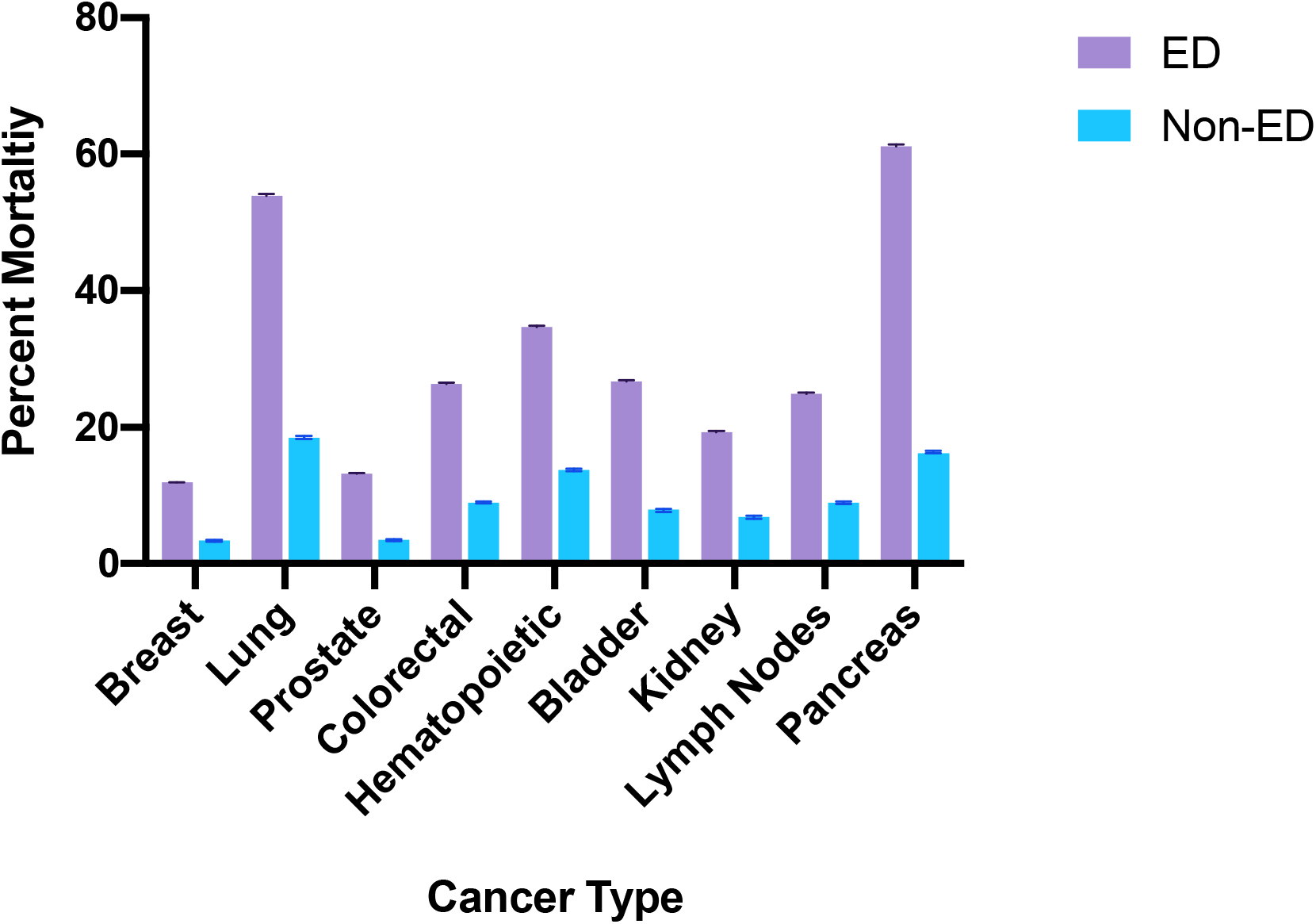
Bar graph comparing mortality percentages from ED-diagnosed to non-ED diagnosed over 5-year study period, error bars representing 95% CI.

**Figure 2.**
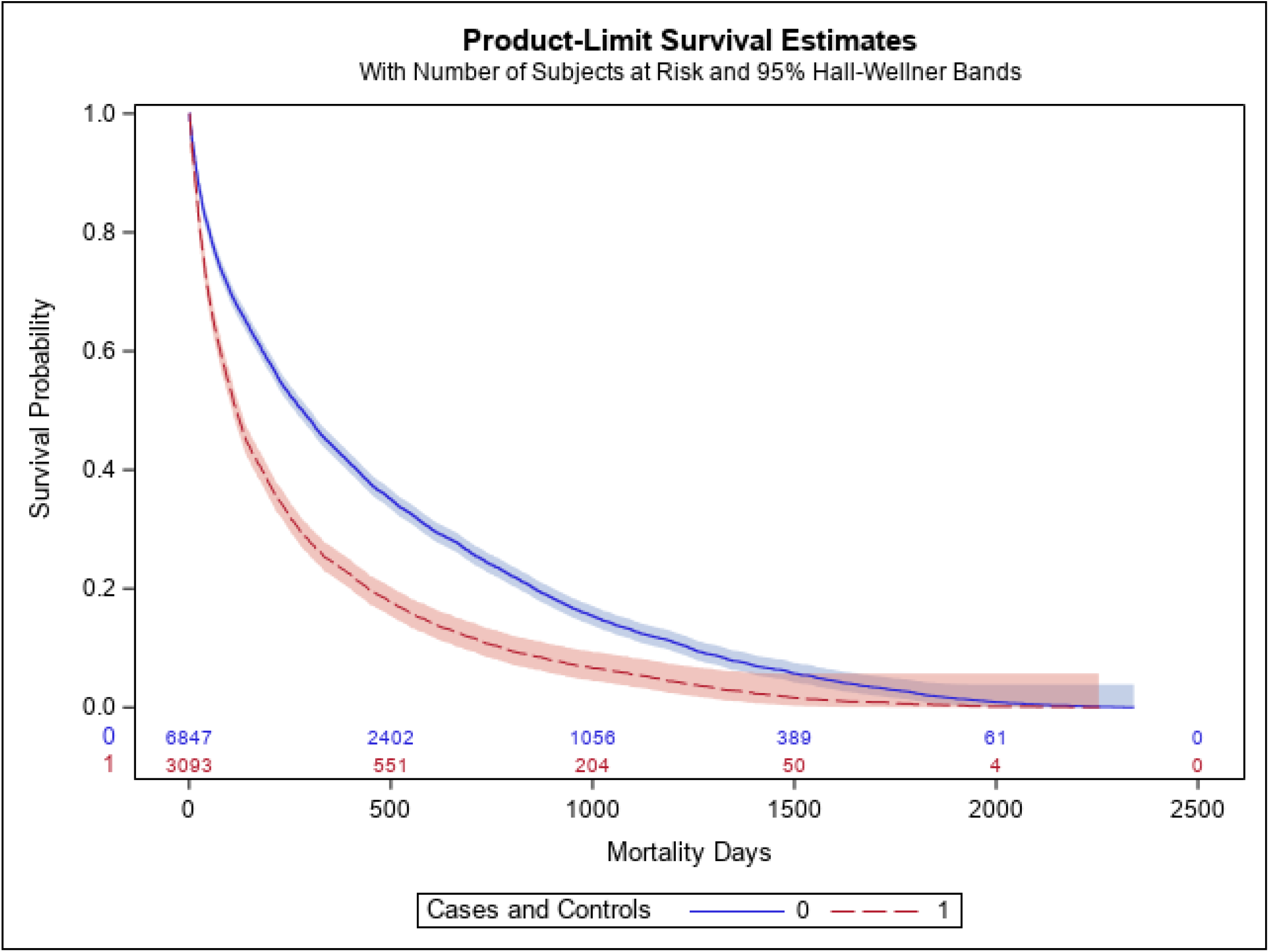
Kaplan Meier survival curve comparing days lived from ED diagnosed to non-ED diagnosed.

### Socioeconomic Status and Racial Outcomes

Table 3 compares SES data between cases and controls, and was assigned by zip code into 5 different categories, namely below/near poverty, low income, middle class, upper middle class, and highest tax bracket.^7,8^ Among cases, 36.9% were low income or below poverty level, compared with 29.5% in the non-ED cohort (95% CI for difference of 7.4% = 0.03-0.04). Supplemental table 3 breaks down the mortality among cases and controls based upon SES. Similar to the data presented in Figures 1 and 2, mortality among cases consistently worse; overall, 36.3% of low-income cases having died in the study period, whereas only 11% low-income controls died.

**Table 3.** Socioeconomic status for controls vs. cases.

| **Total**<br>N=134,761 | **Controls (No ED visit within six months prior) N=118,599**<br>N (total cohort percent, column percent) | **Cases (ED visit within six months prior) N=15,344**<br>N (total cohort percent, column percent) |
| --- | --- | --- |
| **Socioeconomic status\*** |  |  |
| Below or near poverty level | 214 (0.16, 0.18) | 31 (0.02, 0.20) |
| Low income | 34,794 (25.98, 29.34) | 5,583 (4.17, 36.39) |
| Middle class | 83,568 (62.39, 70.46) | 9,729 (7.26, 63.41) |
| Upper middle class | 10 (0.01, 0.01) | 0 (0, 0) |
| Highest tax brackets | 13 (0.01, 0.01) | 1 (0, 0.01) |

\*See text for description of socioeconomic class

Black race consistently worsened cancer prognosis among cases (Supplemental Table 2). For breast cancer, mortality rate was 12.1% for Black patients who were ED cases, versus a mortality rate of 5.5% for Black patients who were in the control group (95% CI for 6.6% difference = 0.01-0.04). Black patients comprised 18.2% of the breast cancer patients among cases versus 8.0% of controls (95% CI for 10.1% difference = 0.06-0.09). These trends continued for lung cancer, with a mortality rate of 58.7% among Black patients in case group, compared with a mortality rate of 22.4% among the control group.

### Controlling for confounders

Table 4 shows the result of logistic regression to examine the effect of case or control status on mortality while controlling for confounding variables. The equation was designed to control for clustering by city (via zip code) and also for seven other potential independent predictor variables (age, gender, race, SES, drug use, alcohol use, tobacco use, and the CCI) and found cases had an increased risk of death over controls with an adjusted odds ratio of 4.12 (95% CI 3.72-4.56). After these corrections, the lower limit of the 95% CI for mortality consistently remained above 1.0 for case versus control status for all cancer.

**Table 4.** Mortality risk based upon recent emergency department (ED) visit, minority and socioeconomic status

|  |  |
| --- | --- |
| Recent ED visit | Odds Ratio (95% CI) * |
| Seen 6 months prior (Unadjusted) | 4.93 (4.40-5.51) |
| Seen 6 months prior (Adjusted**) | 4.12 (3.72-4.56) |
| Minority status | Odds Ratio (95% CI) * |
| Seen 6 months prior (Unadjusted) | 1.55 (1.24-1.94) |
| Seen 6 months prior (Adjusted**) | 0.97 (0.96-0.99) |
| Low socioeconomic status | Odds Ratio (95% CI) * |
| Seen 6 months prior (Unadjusted) | 1.37 (1.01-1.87) |
| Seen 6 months prior (Adjusted**) | 1.19 (0.94-1.50) |
| *Odds of death comparing cases with controls. |  |
| ** Adjusted for age, gender, race, SES, drug use, alcohol use, tobacco use, & Charlson Score. |  |

## Discussion

Our findings show uniformly worsened outcomes of all patients during a period of five years, especially minority and low-income patients, diagnosed with new cancer diagnosed in temporal proximity to an ED visit. These data provide the inference that patients with undiagnosed cancer who rely on the ED as a source of primary medical care will have a worsened outcome compared with patients who have organized medical care.^6^ Prior work from the United Kingdom’s “Routes-to-Diagnosis” conducted by the Public Health England, suggested 23% of newly diagnosed cancer patients presented emergently and survival rates were much lower for those emergent presenters, which is similar to what we discovered in this work.^9^ In the US, a smaller study in Michigan, restricted to lung and colorectal cancer found patients with ED-associated cancer diagnosis had higher CCIs and more advanced stage cancer and were more likely to be Black than those diagnosed without a recent ED visit, but no data on mortality were presented.^10^ It has been demonstrated that emergent presentations of cancer are associated with lower curative rates and treatments, even when compared to cancers diagnosed “electively” (even at the same stage).^11^ The present data in Figure 2, together with prior findings support the inference that patients who have a diagnosis of cancer temporally connected with an ED visit, suffer from a disparity in their odds of survival. From a health services standpoint, potentially modifiable causes for this disparity include lack of access to primary medical care and cancer screening before diagnosis, increased rate of tobacco and alcohol use, worsened comorbidities at the time of diagnosis, and lack of access to specialty cancer care after diagnosis.

This work employed a state-wide assessment of cancer diagnoses in the state of Indiana. From this, numerous patient factors appear to contribute to the observed mortality and worse outcomes, namely race and socioeconomic status. Meanwhile, factors such as age and sex appear to have little relationship between the association of diagnosis and mortality. This is compared to other works where older age (≥85 years old) are 2.5 times more likely to present with an emergent diagnosis of cancer, when compared to a 65–74-year-old cohort.^12^ Those authors conclude that cancer and age are likely to reflect disease specific factors. Further, our work we excluded pediatric patients (<18 years old), and it is well known that more than half of patients that present with *de novo* cancer diagnoses in the ED are emergently diagnosed.^13^

What limited evidence exists from data obtained in the United States, has demonstrated associations between socioeconomic status and the diagnosis of cancer as an emergency. African Americans in one study had increased odds of emergently diagnosed colorectal cancer (AOR 1.5, 95% CI 1.38-1.63) as compared to a similar white cohort.^14^ This disparity not only exists among the diagnosis of cancer but in the primary and secondary preventions among lower SES populations. Colorectal screening, despite its efficacy and recommendations, has been shown to be low among African Americans and those with low SES, demonstrating an opportunity or intervening on this high risk population in the ED.^15^ Furthermore, the low SES population have inequities that result in poor lifestyle choices, some of which (smoking status, diet, physical activity) are preventable and modifiable if this population had access to equitable opportunities. Cancer mortality among this population has an association between mortality and modifiable risk factors, again at current date are not routinely performed in the ED.^16^ Blacks are diagnosed with breast and lung cancer in the cases versus controls and their outcomes appear to be worse. We speculate that blacks are more dependent upon EDs than whites for unscheduled, emergent care, and thus are more likely to present to an ED emergently for their undiagnosed malignacny.^17^ Similar phenomenon can be applied to whites with low SES, where patients of low SES are more reliant on the ED for their care and are more likely to present emergently with their undiagnosed cancer.^18,19^ These data suggest race and socioeconomic status are more important than other factors, such as comorbidities, since the CCI was equal. While not examined in this large data analyses, there is growing evidence that African Americans have more aggressive tumor biology, such as in several breast cancer studies, which can be speculated that this also contributes to these patients presenting more emergently for their undiagnosed cancer.^20^

Of the cancers that have proven screenings that demonstrate success, lung and colorectal cancers do markedly worse when associated with an ED-visit, 54% vs 18% mortality for lung cancer (P<.0001), and 26.4% vs 9% for colorectal cancer (P<.0001). Breast cancer also has a successful screening modality, namely mammography, and suffers from similar poor outcomes when associated with an ED visit, 11.9% mortality vs 3.4%, ED to non-ED associated.^21^ Prostate cancer screening is controversial and recommendations vary by organization and country, regardless, a screening modality is available and those diagnosed with prostate cancer associated with an ED visit have higher mortality than those that don’t (13.2% vs 3.5%).^22^ Lastly, cervical cancer also is frequently screened for as outpatients and 22% of those seen in the ED were dead, versus 7.4% of those not seen in the ED. This evidence is supported by known disparities in cancer screening, with minority patients experiencing greater delays in evaluation and screening for cancer, leading to suboptimal treatment among those patients subsequently diagnosed with cancer.^23^

Previous research has called for both improving the outcomes of patients that are diagnosed with cancer through an emergent presentation, as well as helping to reduce the burden of emergent diagnoses by improving cancer screening.^6,24^ This is likely a systems issue, but plausible future steps is utilizing the ED space for more than just emergent care. Average length of stays in ED has multiple variables that impact exact time frame which patients sit in the ED, but in one paper an average a patient can expect a wait of 4 hours.^25^ As the trend of increasing length of stay continues to increase nationwide, EDs are experiencing the unfortunate phenomena of ED crowding which has been well demonstrated to be associated with increased hospital death.^26^ We propose future work in developing interventions for this at risk population while in the ED, similar to what has been performed for rapid hepatitis screening and cervical cancer screening in urban EDs, demonstrating proof of concept and utilizing the ED space for more than emergent care.^27,28^ Similar revolutionary changes in ED workflow for improving the overall health of ED patients has been adopted with universal HIV screening in the ED, as well as universal suicide screening in EDs.^29,30^ Intervening on this population is challenging but supported by a Cochrane Review, guaiac fecal immunochemical test (FIT) can reduce colorectal mortality by 15%, and providing appropriately chosen patients with home use FIT tests through primary care has improved screening rates, sustaining a screening rate of 75%.^31,32^ Currently screening routinely for cancer does not occur in the emergency setting, but for many vulnerable patients (uninsured, lower SES, racial minorities), the emergency room serves as the only opportunity for routine care and we should begin to explore alternative strategies to maximally improve the care of these patients.^33^ Removing the barriers to cancer screening, such as providing patients with FIT cards prior to discharge may represent an opportunity to increase adherence to CRC screening and reduce the burden of emergently diagnosed CRC.^34^ Novel approaches need be undertaken at the systems and health policy level to address the disparities that are well demonstrated among ED-associated cancer diagnoses.

### Limitations

There are limitations to this study, namely the retrospective methodology to obtaining the administrative data. A diagnosis of cancer, while suspected in the ED, usually requires a biopsy and extensive workup (PET scan for example), which come weeks to months after the actual ED encounter, and thus confirming linkage from a suspected ED visit and a diagnosis is challenging. Further, patient stage and tumor specific biology are not included on the ICD-coded diagnosis and thus knowing the stage and extent of disease is not available without retrospective chart review. Tumor specific biology has been linked to increased associations with emergent presentations, with more rapidly, aggressive tumors being more likely to be diagnosed emergently.^35^ Regardless, we made an inference that being diagnosed with cancer within 6 months of a recorded ED visit, meant those ED physicians had an opportunity to diagnose asymptomatic cancer, if that ED visit wasn’t directly related to the presentation of emergent cancer diagnosis. Further, no knowledge is known about previous screenings and primary care follow up, thus no inference can be made to know whether or not screening may have reduced the likelihood of the found associations. Further limitations include those cancers that are so advanced that tissue biopsy is not obtained, or patients prefer to not seek treatment. The lack of follow-up knowledge means we can’t examine the relationship between mortality and inadequate access to expert care, but the correction for clustering on logistic analyses suggests this is not just a result of location.

A last limitation and that of an area of future research is exploration into pediatric cancers and their association with being emergently diagnosed in the ED. No pediatric cancers have preventable or modifiable risk factors, and as such the goal of this work is to find interventions on cancers that can be prevented with lifestyle modification (smoking cessation, weight loss), or caught earlier with age-appropriate screening.

## Conclusions

Patients diagnosed with cancer with an associated ED visit within 6 months prior to their ICD-code cancer diagnosis are associated with poor outcomes, specifically increased mortality, as compared to a cohort that did not have an ED visit within 6 months prior to their diagnosis. Lung, breast, and colorectal are the most frequently ED-associated cancer diagnoses, with upwards of 50% mortality as compared to non-ED associated cancers. Further, there are associated racial and socioeconomic disparities among those diagnosed, both in cancer type and frequency, and mortality. These data are among the first to our knowledge that describe patients in the United States, among a statewide database. They further suggest that ED-associated cancer diagnoses offers an opportunity for additional research to understand the associations between diagnoses and socioeconomic and racial disparities. Given existing literature is limited to retrospective, database analyses, future work should be aimed at prospective studies as to guide future interventions to help reduce the disparities and reduce the mortality among ED-associated cancer diagnoses.

## Data Availability

Data available upon request

## Abbreviations

ED: emergency department
CCI: Charlson Comorbidity Index

---

**Supplemental table 1.**
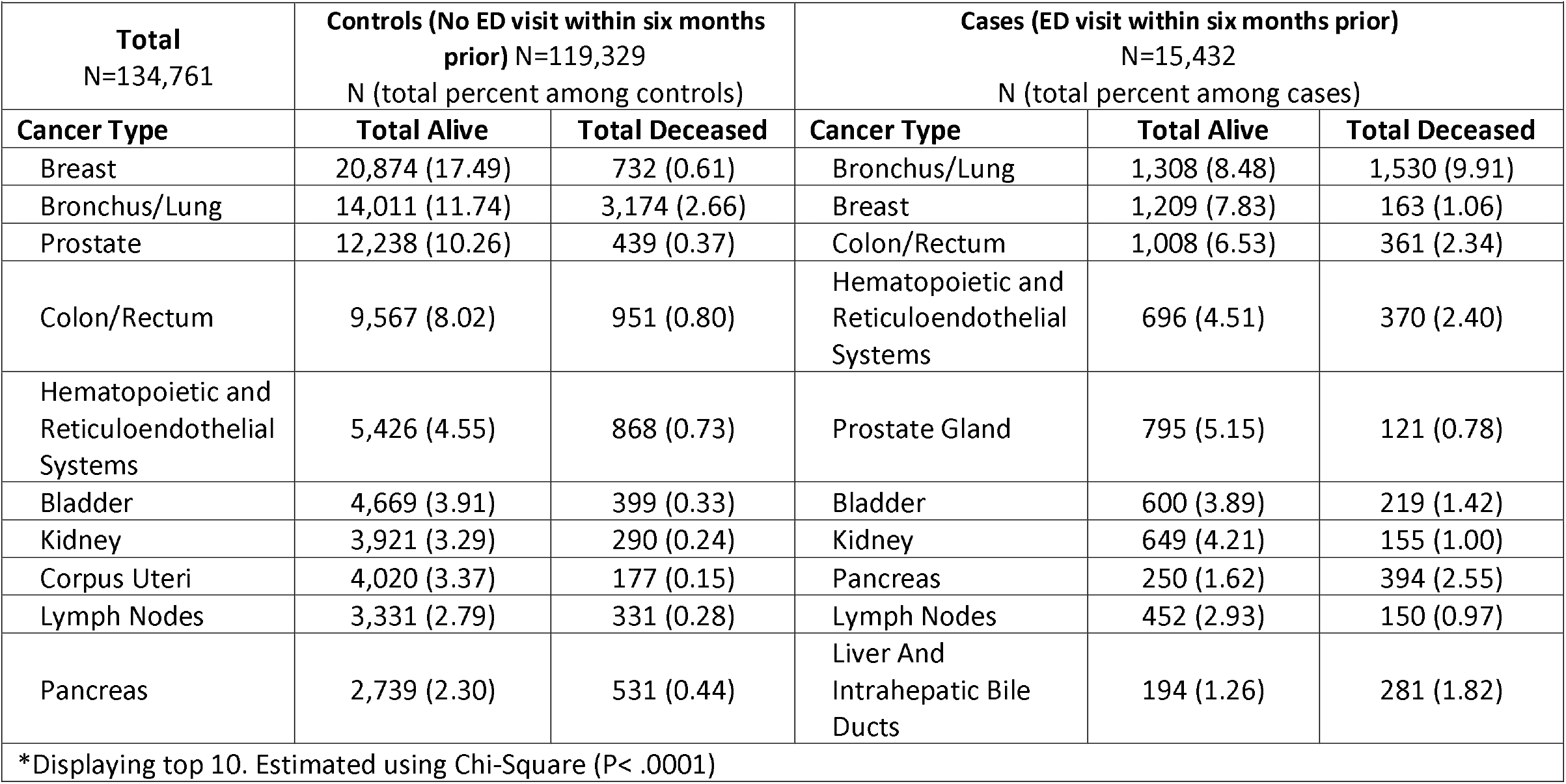
Mortality frequency by cancer type for controls vs. cases.

**Supplementary Table 2.**
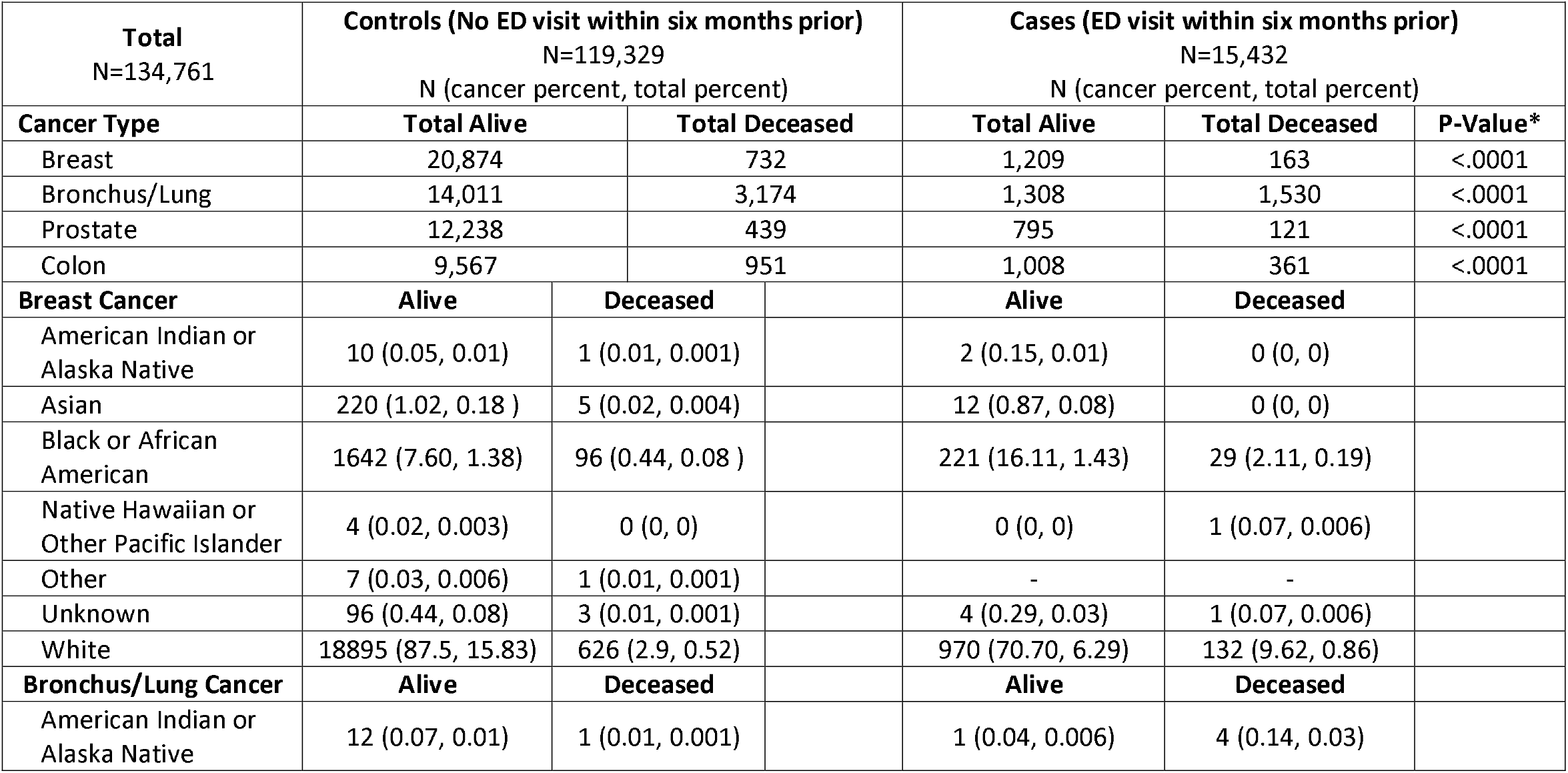
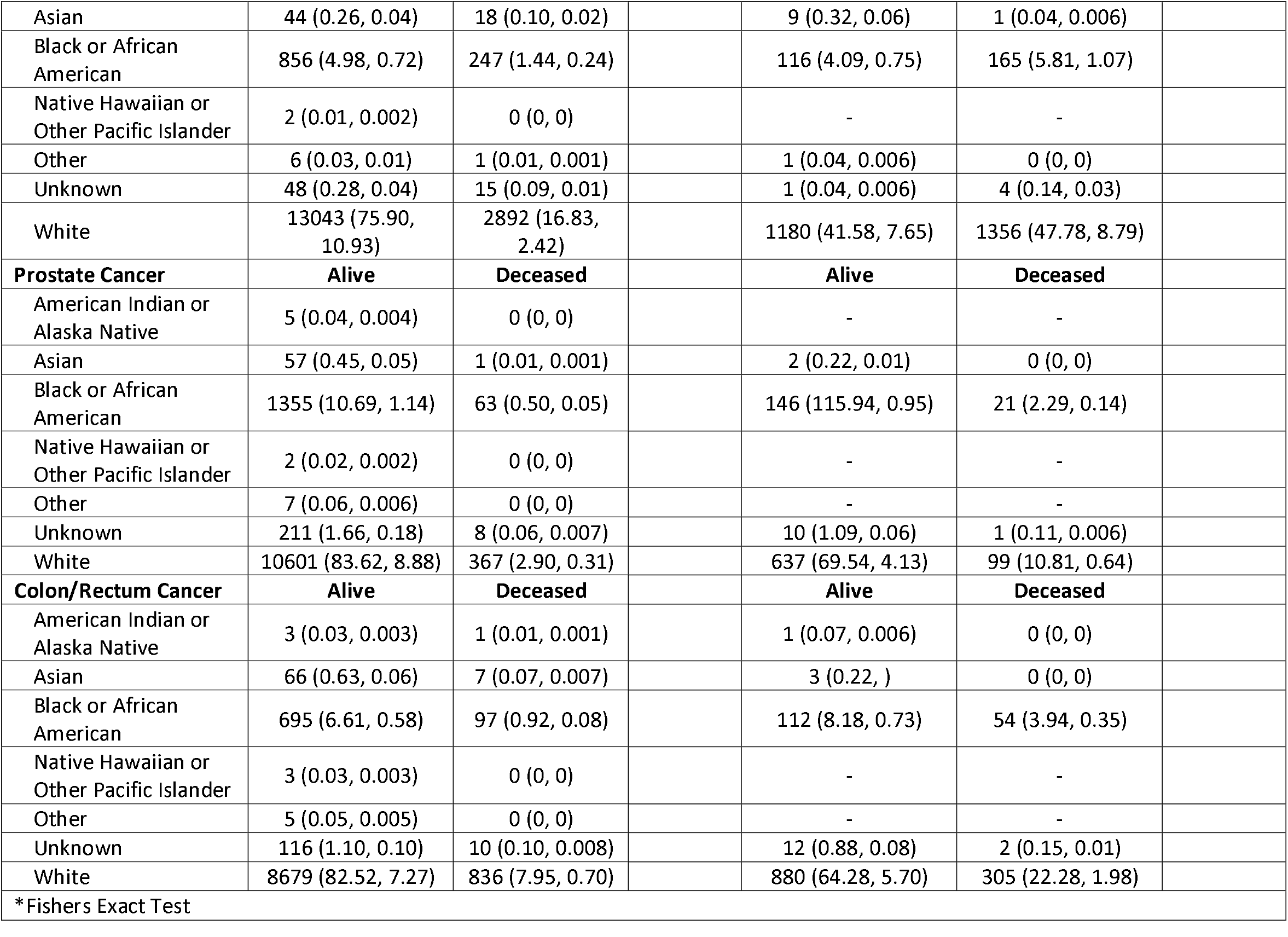
Mortality frequency for top 4 cancers, by race and cancer type.

**Supplemental Table 3.**
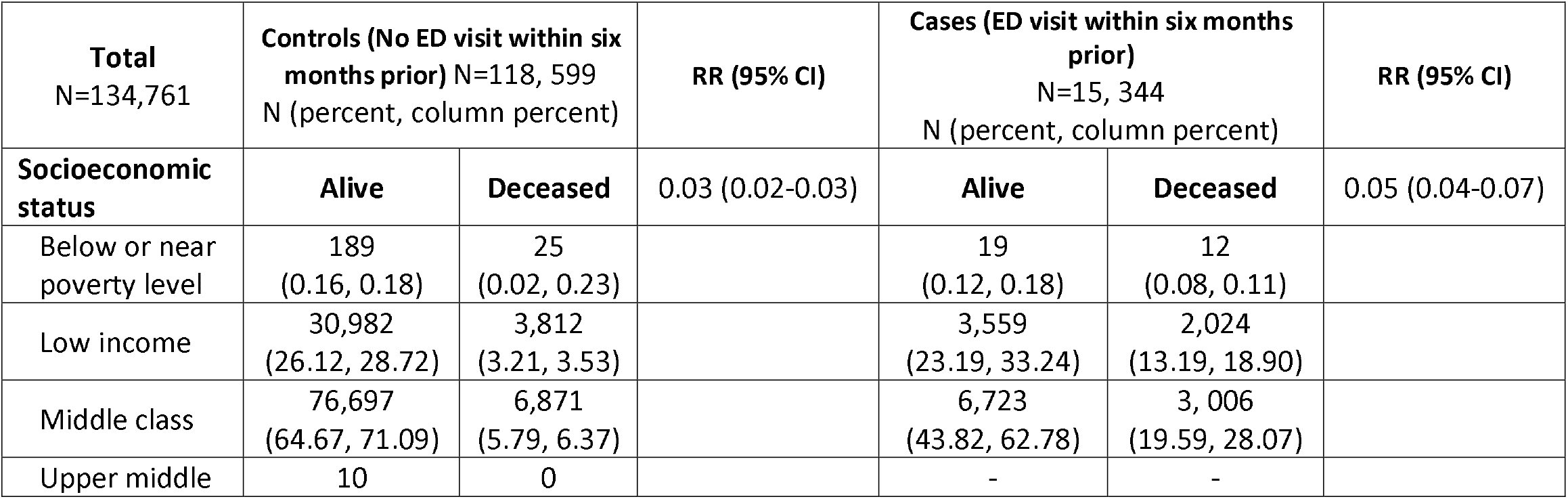
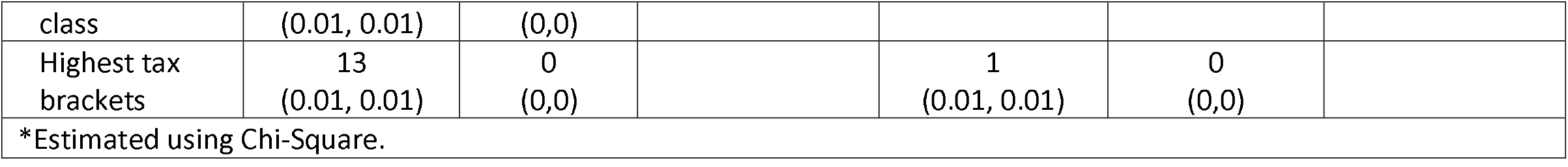
Mortality between cases and controls, stratified by income.

## Notes

### Competing Interest Statement

The authors have declared no competing interest.

### Funding Statement

No external funding was received

### Author Declarations

IU IRB approved under exempt status, protocol #2004425945

## References

1. Heron M, Anderson RN. Changes in the Leading Cause of Death: Recent Patterns in Heart Disease and Cancer Mortality. NCHS Data Brief. 2016(254):1–8.

2. Miller KD, Nogueira L, Mariotto AB, et al. Cancer treatment and survivorship statistics, 2019. CA Cancer J Clin. 2019;69(5):363–385.

3. Khorana AA, Tullio K, Elson P, et al. Time to initial cancer treatment in the United States and association with survival over time: An observational study. PLoS One. 2019;14(3):e0213209.

4. Richards TB, Doria-Rose VP, Soman A, et al. Lung Cancer Screening Inconsistent With U.S. Preventive Services Task Force Recommendations. Am J Prev Med. 2019;56(1):66– 73.

5. Schwartz LM, Woloshin S, Fowler FJ, Jr., Welch HG. Enthusiasm for cancer screening in the United States. JAMA. 2004;291(1):71–78.

6. Zhou Y, Abel GA, Hamilton W, et al. Diagnosis of cancer as an emergency: a critical review of current evidence. Nat Rev Clin Oncol. 2017;14(1):45–56.

7. Bureau USC. American Community Survey.

8. CUP) HCaUPH. NIH Description of Data Elements. 2008.

9. Service NCRaA. Routes to Diagnosis - NCIN Data Briefing. 2010.

10. Sikka V, Ornato JP. Cancer diagnosis and outcomes in Michigan EDs vs other settings. Am J Emerg Med. 2012;30(2):283–292.

11. Gunnarsson H, Jennische K, Forssell S, et al. Heterogeneity of colon cancer patients reported as emergencies. World J Surg. 2014;38(7):1819–1826.

12. Abel GA, Shelton J, Johnson S, Elliss-Brookes L, Lyratzopoulos G. Cancer-specific variation in emergency presentation by sex, age and deprivation across 27 common and rarer cancers. Br J Cancer. 2015;112 Suppl 1:S129–136.

13. Elliss-Brookes L, McPhail S, Ives A, et al. Routes to diagnosis for cancer - determining the patient journey using multiple routine data sets. Br J Cancer. 2012;107(8):1220–1226.

14. Pruitt SL, Davidson NO, Gupta S, Yan Y, Schootman M. Missed opportunities: racial and neighborhood socioeconomic disparities in emergency colorectal cancer diagnosis and surgery. BMC Cancer. 2014;14:927.

15. Warren Andersen S, Blot WJ, Lipworth L, Steinwandel M, Murff HJ, Zheng W. Association of Race and Socioeconomic Status With Colorectal Cancer Screening, Colorectal Cancer Risk, and Mortality in Southern US Adults. JAMA Netw Open. 2019;2(12):e1917995.

16. Hastert TA, Ruterbusch JJ, Beresford SA, Sheppard L, White E. Contribution of health behaviors to the association between area-level socioeconomic status and cancer mortality. Soc Sci Med. 2016;148:52–58.

17. Blanchard JC, Haywood YC, Scott C. Racial and ethnic disparities in health: an emergency medicine perspective. Acad Emerg Med. 2003;10(11):1289–1293.

18. Krieg C, Hudon C, Chouinard MC, Dufour I. Individual predictors of frequent emergency department use: a scoping review. BMC Health Serv Res. 2016;16(1):594.

19. Wachelder JJH, van Drunen I, Stassen PM, et al. Association of socioeconomic status with outcomes in older adult community-dwelling patients after visiting the emergency department: a retrospective cohort study. BMJ Open. 2017;7(12):e019318.

20. Keenan T, Moy B, Mroz EA, et al. Comparison of the Genomic Landscape Between Primary Breast Cancer in African American Versus White Women and the Association of Racial Differences With Tumor Recurrence. J Clin Oncol. 2015;33(31):3621–3627.

21. Caughran J, Braun TM, Breslin TM, et al. The Effect of the 2009 USPSTF breast cancer screening recommendations on breast cancer in Michigan: A longitudinal study. Breast J. 2018;24(5):730–737.

22. Kim EH, Andriole GL. Prostate-specific antigen-based screening: controversy and guidelines. BMC Med. 2015;13:61.

23. Fiscella K, Humiston S, Hendren S, et al. Eliminating disparities in cancer screening and follow-up of abnormal results: what will it take? J Health Care Poor Underserved. 2011;22(1):83–100.

24. Goodyear SJ, Leung E, Menon A, Pedamallu S, Williams N, Wong LS. The effects of population-based faecal occult blood test screening upon emergency colorectal cancer admissions in Coventry and north Warwickshire. Gut. 2008;57(2):218–222.

25. Mentzoni I, Bogstrand ST, Faiz KW. Emergency department crowding and length of stay before and after an increased catchment area. BMC Health Serv Res. 2019;19(1):506.

26. Boulain T, Malet A, Maitre O. Association between long boarding time in the emergency department and hospital mortality: a single-center propensity score-based analysis. Intern Emerg Med. 2019.

27. White DA, Anderson ES, Pfeil SK, Trivedi TK, Alter HJ. Results of a Rapid Hepatitis C Virus Screening and Diagnostic Testing Program in an Urban Emergency Department. Ann Emerg Med. 2016;67(1):119–128.

28. Hogness CG, Engelstad LP, Linck LM, Schorr KA. Cervical cancer screening in an urban emergency department. Ann Emerg Med. 1992;21(8):933–939.

29. Orkin C, Flanagan S, Wallis E, et al. Incorporating HIV/hepatitis B virus/hepatitis C virus combined testing into routine blood tests in nine UK Emergency Departments: the “Going Viral” campaign. HIV Med. 2016;17(3):222–230.

30. Heyland M, Delaney KR, Shattell M. Steps to Achieve Universal Suicide Screening in Emergency Departments: A Call to Action. J Psychosoc Nurs Ment Health Serv. 2018;56(10):21–26.

31. Bakhai S, Ahluwalia G, Nallapeta N, Mangat A, Reynolds JL. Faecal immunochemical testing implementation to increase colorectal cancer screening in primary care. BMJ Open Qual. 2018;7(4):e000400.

32. Nicholson BD, Thompson M, Price CP, Heneghan C, Pluddemann A. Home-use faecal immunochemical testing: primary care diagnostic technology update. Br J Gen Pract. 2015;65(632):156–158.

33. Begley CE, Vojvodic RW, Seo M, Burau K. Emergency room use and access to primary care: evidence from Houston, Texas. J Health Care Poor Underserved. 2006;17(3):610– 624.

34. Cusumano VT, May FP. Making FIT Count: Maximizing Appropriate Use of the Fecal Immunochemical Test for Colorectal Cancer Screening Programs. J Gen Intern Med. 2020;35(6):1870–1874.

35. Comber H, Sharp L, de Camargo Cancela M, Haase T, Johnson H, Pratschke J. Causes and outcomes of emergency presentation of rectal cancer. Int J Cancer. 2016;139(5):1031– 1039.

